# Classification of the infection status of COVID-19 in 190 countries

**DOI:** 10.1101/2020.12.17.20248445

**Authors:** Takashi Odagaki, Reiji Suda

**Affiliations:** Kyushu University, Nishiku, Fukuoka 819-0395, Japan; Research Institute for Science Education, Inc., Kitaku, Kyoto 603-8346, Japan

## Abstract

We propose a simple method to determine the infection rate from the time dependence of the daily confirmed new cases, in which the logarithm of the rate is fitted by piece-wise quadratic functions. Exploiting this method, we analyze the time dependence of the outbreak of COVID-19 in 190 countries around the world and determine the status of the outbreak in each country by the dependence of the infection rate on the number of new cases. We show that the infection status of each country can be completely classified into nine different states and that the infection status of countries succeeded in controlling COVID-19 implies the importance of the quarantine and/or self-isolation measure.

## Introduction

The pandemic COVID-19 appears to have entered a new phase in the winter of 2021 after the upward trend in the fall of 2020 followed by the steep downward trend in many countries. The time dependence of daily confirmed new cases in each country, which we call an infection curve for simplicity, shows different behavior including a wavy curve with different number of peaks [1]. In order to formulate policies against COVID-19 in each country, it is desirable to have a standard classification scheme by which the status of the outbreak of COVID-19 can be characterized. Furthermore, the classification scheme must necessarily be based on limited information such as the infection curve.

Conventionally, the outbreak of an epidemic is understood by mean field approaches in which the population are separated into several compartments. The COVID-19 is different from known epidemics in that there are many asymptomatic infected individuals who are infectious, pre-symptomatic patients are also infectious and the PCR test can be used to identify infected individuals. Therefore, the minimal compartments to understand COVID-19 are individuals (R) immune to SARS-CoV-2 by recovery from the infection and/or by vaccination, susceptible individuals (S), infectious infected-individuals in the community (I) and non-infectious infected-individuals (Q) who are quarantined or self-isolated at home or in some other places after tested positive. Since there are many asymptomatic infected-individuals, the number of individuals in compartment I cannot be known. However, if the PCR test is conducted properly and the individuals tested positive comply strictly with the regulations, the number of individuals in compartment Q, or the daily confirmed new cases Δ *Q*, can be treated as an observable of COVID-19 which can be used in a theoretical analysis.

In this paper, we propose a simple method to approximate the infection curve and to obtain the rate of change of the number of daily confirmed new cases, which we call the infection rate for simplicity. We show that the relation between the infection rate and the number of daily confirmed new cases can be used to define the status of outbreak and that there are nine qualitatively distinguishable states in the evolution of the outbreak. Namely, the infection status can be characterized as T-*k*_*n*_ where T denotes one of four trends (increasing (I) or the epidemic rise, stationary (S) or the plateau, decreasing (D) or the epidemic slow down and converged (C) or virtually no infected individuals), *k* and *n* represent one of six stages and the number of repetitions (waves) of infection status, respectively. We analyze the infection curve in 190 countries and classify them into the nine states. It is important to note that the present analysis can be used in any stage of outbreak including the stage of the herd immunity.

We first explain the fitting procedure for the infection curve and analyze the infection status of ten typical countries. Then, we discuss the evolution of infection status in the plot of the infection rate against the number of the daily confirmed new cases, which is called an infection status plot, and show that the infection status can be classified into nine states. We analyze the time dependence of the daily confirmed new cases of 190 countries and classify the infection status of these countries. Results are discussed at the end.

### Procedure for fitting the infection curve

The infection curve shows a wavy time dependence with series of maxima and minima when it is fitted by a continuous function. We assume that the function can be separated into basic units each of which consists of inflection, maximal, inflection, minimal and inflection points. We denote the time of these breaking points of the *i*-th unit as *t* _4*i* −_ 4, *t*_4*i* −_ 3, *t*_4*i* −_ 2, *t*_4*i* −_ 1 and *t*_4*i*_, The upper panel of Fig. 1 shows these points for the first unit.

**Figure 1:**
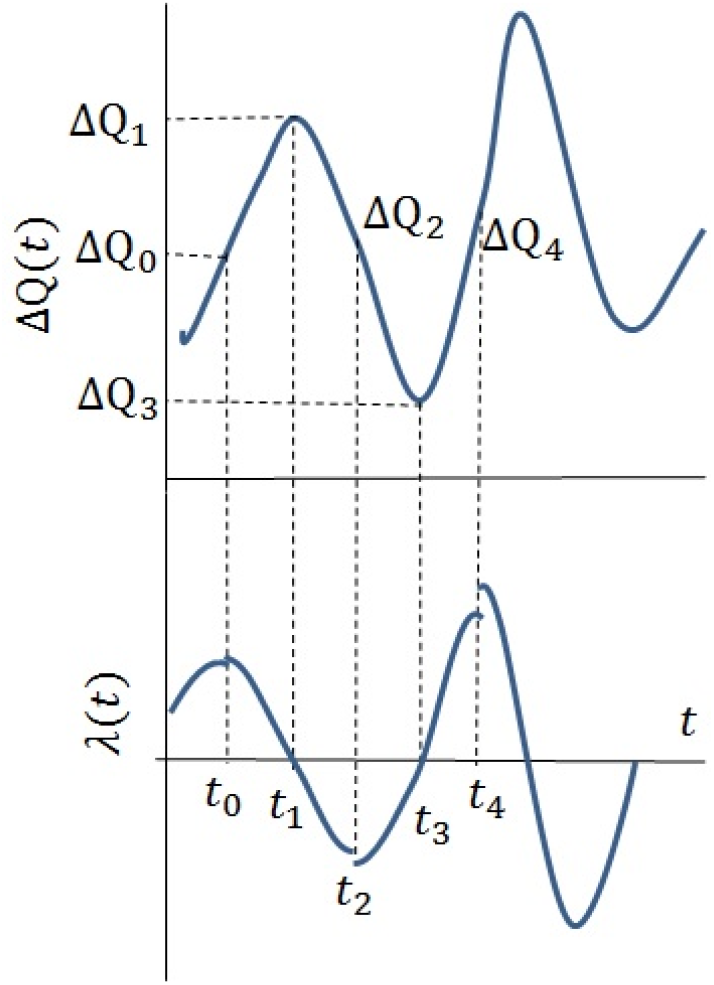
Setting of parameters for an infection curve. In each time period, *λ(t)* is approximated by a quadratic function shown in Table 1.

We fit Δ *Q (t)* by piece-wise continuous functions which are determined between an inflection point and its adjacent extremum point. To this end, we express Δ *Q (t)* in the region between two breaking points at *t* _*m*_ and *t*_*n*_ (*t*_*m*_ < *t*_*n*_)

**Table 1:** Piece-wise functions for fitting of a unit shown in Fig. 1.

| Period | $\lambda(t)$ | $\Delta Q(t)$ |
| --- | --- | --- |
| $t_0 \sim t_1$ | $f(\alpha_0, t_0, t_1, t)$ | $\Delta Q_0 \exp\{g_1(\alpha_0, t_0, t_1, t)\}$ |
| $t_1 \sim t_2$ | $f(\alpha_1, t_2, t_1, t)$ | $\Delta Q_1 \exp\{g_2(\alpha_1, t_2, t_1, t)\}$ |
| $t_2 \sim t_3$ | $f(\alpha_2, t_2, t_3, t)$ | $\Delta Q_2 \exp\{g_1(\alpha_2, t_2, t_3, t)\}$ |
| $t_3 \sim t_4$ | $f(\alpha_3, t_4, t_3, t)$ | $\Delta Q_3 \exp\{g_2(\alpha_3, t_4, t_3, t)\}$ |

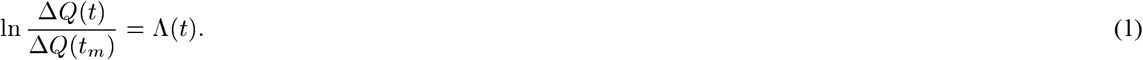

In order to make the argument transparent, we define the infection rate *λ (t)* by

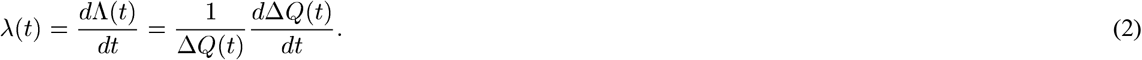

As shown in the lower panel of Fig. 1, it is apparent that *λ (t)* is zero when Δ *Q (t)* takes its extrema, namely

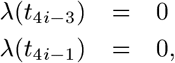

and that *λ (t)* becomes a maximum at the inflection point at *t*_4*i*−_4 and a minimum at the inflection point at *t*_4*i*−_2. These properties of *λ (t)* allow us to approximate it in each section by a quadratic function *λ (t) = f (a, b, c, t)* where

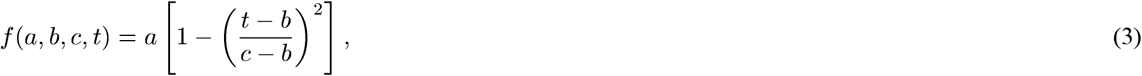

It is easy to check that *f (a, b, c, c) =* 0 and *f (a, b, c, b) = a* is the extremum of *f (a, b, c, t)*.

With the piecewise approximation of *λ (t)*, it is straight forward to determine the piecewise approximation of Λ *(t)* or Δ *Q (t) = (t*_*m*_) exp[Λ *(t)*]. We find that Λ *(t) = g(a, b, c, t)* where *g(a, b, c, t)* is given by

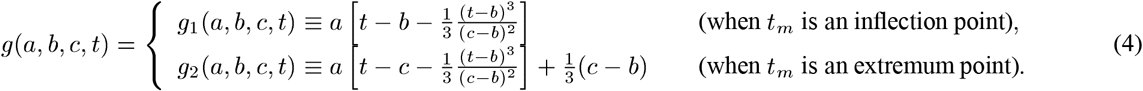

Table 1 summarizes the fitting procedure for the first unit, where *t* _*j*_’s (*j = 0, 1, 2, 3, 4*) are the breaking points and the values of Δ *Q (t)* at these points are denoted as Δ *Q* _*j*_ *=* Δ *Q (t*_*j*_). Here, *α* _*j*_’s are the value of *λ (t)* at its extrema.

In order to determine *α* _*i*_’s, we impose the continuity of Δ *Q (t)* at *t*_*j*_’s and find that

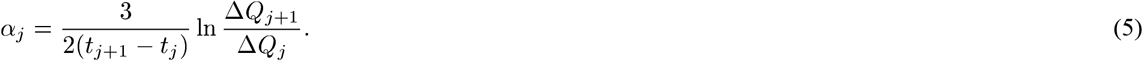

Note that *α* _*j*_ and *α* _*j*+1_ may not be the same since we do not requite the continuity of *λ (t)* and allow that the piecewise functions for Δ *Q (t)* may have different slopes on each side of the inflection point. Although this does not affect the analysis of the infection curve and the infection status plot, it might be reflection of an abrupt change in policies.

Figure 2 shows the analysis of the infection curve in 10 typical countries from January 23, 2020 to February 17, 2021. The data are taken from Coronavirus Resource Center, Johns Hopkins University [1]. Since the time dependence of the infection rate and the number of new cases are given by explicit functions, we plot *λ* as a function of Δ *Q* (the infection status plot) which are also shown in Fig. 2.

**Figure 2:**
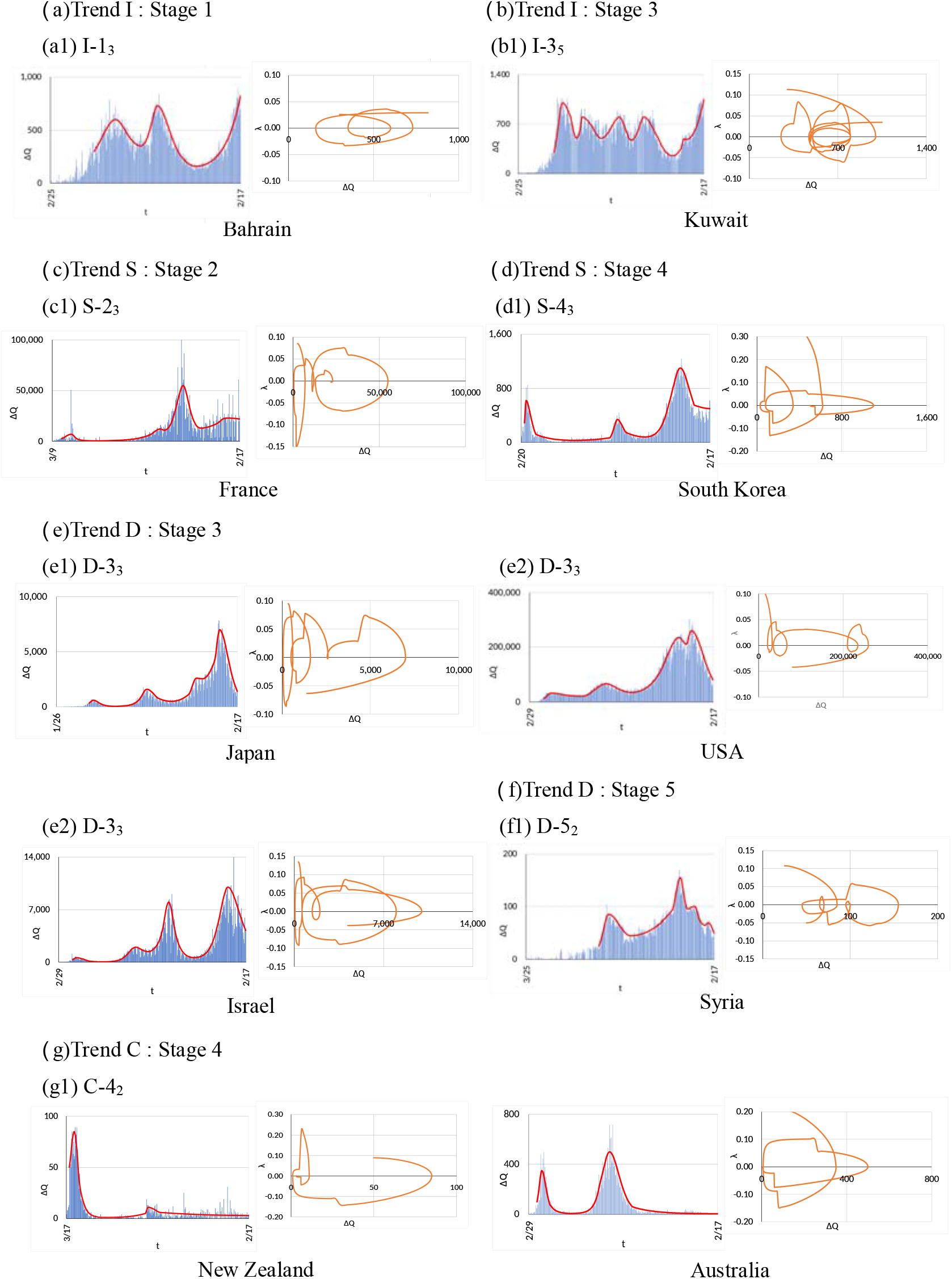
Infection curves from January 23, 2020 to February 17, 2021 for ten countries in different trends: (a) and (b) increasing trend, (c) and (d) stationary trend, (e) and (f) decreasing trend and (g) converged. See the following section for the classification scheme. In each country, the panel on the left hand side is the infection curve Δ *Q (t)* with fitting by Eq. (1), and the panel on the right hand side in each country is the infection status plot, *λ* vs Δ *Q*. Note differences in the scale of axes. Data from Coronavirus Resource Center, Johns Hopkins University [1].

We have analyzed the infection curves of 190 countries in the same period, 24 typical samples of which are shown in Fig. 1 in the supplementary materials. As can be seen from these analyses, the infection status is different from country to country and seems to be classified by the trends, which will be discussed in the following section.

### Infection statuses of 190 countries

#### Evolution of infection status

Since the first outbreak of COVID-19, the infection situation has been evolving in each countries. In order to establish a classification scheme, we first define the trends and stages of the infection status by the following procedure:

1. We distinguish four trends of the infection curve at a given instance by the sign of *λ* ; increasing (I: *λ* > 0), stationary (S: *λ =* 0), decreasing (D: *λ* < 0) and converged (C: *λ* ≤ 0 and Δ *Q* ≅ 0) states. In the converged state, there might be intermittent appearance of infected individuals, but they are controlled immediately.
2. Stationary states or the plateau of the infection curve are considered to constitute stages. The trajectory between adjacent stationary states is either increasing or decreasing, which belong to different stages. The stages are numbered by the time sequence.
3. When the trajectory passes *λ =* 0 line from the lower half-plane to upper half-plane for *n* times, the current status is considered to be in the (*n* + 1)th wave. However, small loops of the trajectory around *λ =* 0 are not included as waves.
4. When the trajectory goes from *λ =* 0 to the upper or lower half-plane, the corresponding stage is kept until the trajectory becomes horizontal.

Following this procedure, we summarize the evolution of the infection status as shown in Fig. 3. The outbreak of COVID-19 starts from S-0 in any countries. While some countries stay in this stage with a few patients appearing intermittently, the infection expands in most of the countries moving to Stage 1 of Trend I. At Stage 1, the epidemic will either keep increasing (I-1 state) or reach a maximal stationary state (S-2 state) and may stay there for a long time until the herd immunity is realized. The state of S-2 starts either increasing again (I-3 state) or decreasing (D-3 state) before the herd immunity becomes effective. State I-3 will shift to state I-1 and state D-3 will move either to converged state C-4 or to stationary state S-4 corresponding to a minimal point of Δ *Q (t)*. From the minimal point, it can expand again (I-5 state) or decrease again (D-5 state) at Stage 5. The infection status in I-5 state goes back to equivalent I-1 state, and the infection status D-5 goes back to equivalent D-3 state. Therefore, we expect that there are nine qualitatively different states in the outbreak of COVID-19.

**Figure 3:**
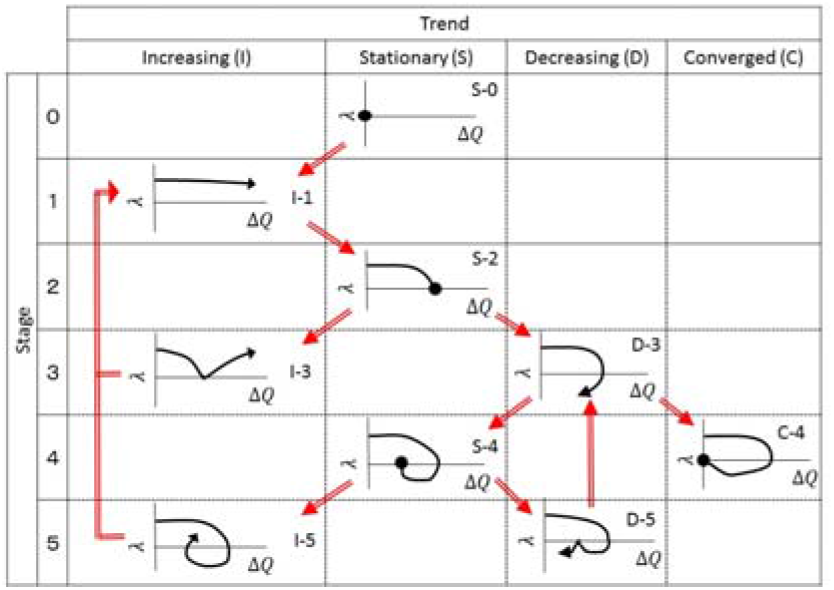
Classification of infection status by the infection status plot. Solid circles denote that the plot is stationary. Red arrows indicate the direction of the possible change of the states. I-3 and I-5 are equivalent to I-1, and D-5 is equivalent to D-3.

In Fig. 4, we show schematically infection curves corresponding to Fig. 3.

**Figure 4:**
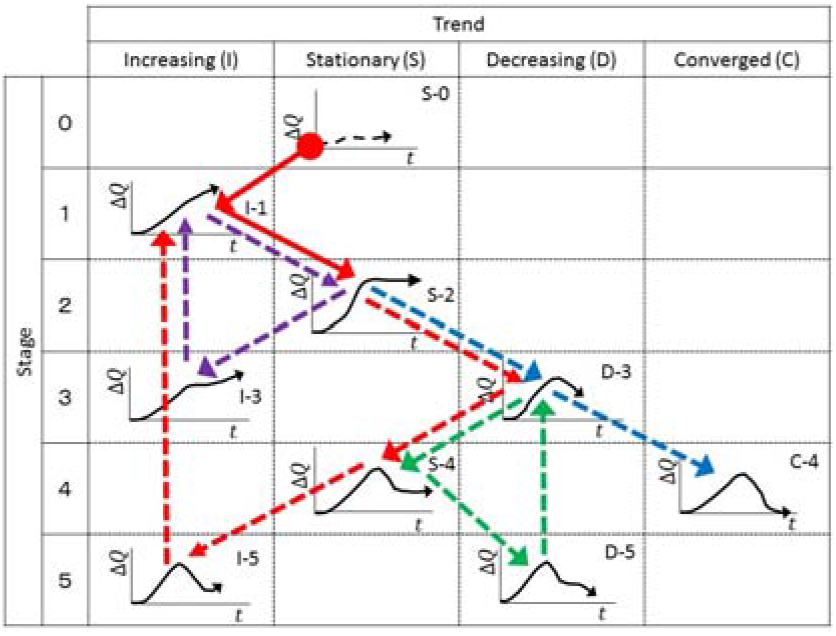
The infection curves for different infection states. The solid red arrows indicate the process of outbreak in most of the countries. Red dashed-arrows are the trajectory of a wavy infection and blue dashed-arrows indicate an efficient conversion process. Step-wise increase and step-wise decrease are denoted by purple dashed-arrows and green dashed-arrows, respectively.

We show typical trajectories by colored arrows in Fig. 4. We find three possible repeated evolutions of infection in Figs. 3 and 4. One repeated infection is I-1 → S-2 → D-3 → S-4 → I-5→ I-1, which is shown by a red arrow and red dashed-arrows in Fig. 4. This wavy infection has been observed in many countries such as Japan, USA and Kuwait. The center position of the second round and subsequent loop of the trajectory in the infection status plot may drift horizontally in either directions. One of other two repeated behaviors is I-1 → S-2 → I-3 → I -1 as shown by purple dashed-arrows in Fig.4, which represents a step-wise increase of as seen in Morocco and Lebanon (See Fig. 3 in the supplementary materials). The other is D-3 → S-4 → D-5 → D-3 as shown by green dashed-arrows in Fig. 4, which indicates a step-wise decrease of Δ *Q* as seen in Philippines (See Fig. 4 in the supplementary materials). The trajectory S-2 → D-3 → C-4 as shown by blue dashed-arrows represents the most efficient control of COVID-19 by strong measures seen in China, Taiwan and some other countries.

### Classification of 190 countries

Using the methodology and definitions described in the previous subsection, we classified the statuses of the COVID-19 outbreaks in 190 countries as of February 17, 2021 into nine states. Results are summarized in Table 2. In the supplementary materials, we show similar analyses as of November 19, 2020. It is apparent that the evolution of the infection status depends strongly on the policies of each country. Countries succeeded in controlling COVID-19 used a strong measure on social distancing and on quarantining and strict control of travelers from abroad. On the other hand, countries suffering several waves like Japan, USA and many other countries have lifted measures in state S-4 putting much weight on corona economic measures and have brought into the state of *λ* > 0 again. It has been shown that the wavy infection curve is self-organized by such a policy [2, 3].

**Table 2:** Classification of the infection status of COVID-19 in 190 countries as of February 17, 2021. The first and second columns are Trend and Stage, respectively. The infection status is identified by Trend, Stage and Wave defined in the previous subsection and the status is labeled as T-*k*_*n*_, where T denotes a trend (increasing (I), stationary (S), decreasing (D) and converged(C)), *k* and *n* represent a stage and the current wave. Countries are listed in alphabetical order in each state.

| T | S | Countries |  |  |
| --- | --- | --- | --- | --- |
|  |  | Wave 1 | Wave 2 | Wave 3 ~ |
| I | 1 | Antigua and Barbuda, Saint Lucia | Benin, Gabon, Gambia, Guinea-Bissau, Sao Tome and Principe, South Sudan | Bahrain, Jamaica, Somalia |
|  | 3 | Barbados, Seychelles | Mongolia, Sri Lanka | Kuwait, Maldives, Montenegro |
|  | 5 | Ethiopia, Greece, Hungary, Iraq, Jordan, Moldova, Qatar | Bulgaria, Burundi, Czechia, Egypt, Estonia, Namibia, West Bank and Gaza | Iran, Paraguay, Peru |
| S | 0 | Cambodia, Dominica, Fiji, Grenada, Laos, Marshall Islands, Micronesia, Saint Kitts and Nevis, Samoa, Solomon Islands, Timor-Leste, Vanuatu |  |  |
|  | 2 | Botswana, Saint Vincent and the Grenadines | Brazil, Chile, Cote d'Ivoire, Cuba, Ecuador, Mozambique, San Marino, Senegal | Finland, France, Malta, Togo |
|  | 4 | Bahamas, Belize, Bhutan, Bosnia and Herzegovina, Cameroon, Congo (Brazzaville), Croatia, Cyprus, Georgia, Guatemala, Guinea, Iceland, India, Lithuania, Madagascar, Morocco, Nepal, Nicaragua, Papua New Guinea, Philippines, Poland, Romania, Trinidad and Tobago, Ukraine, Uruguay | Afghanistan, Armenia, Austria, Azerbaijan, Bangladesh, Belgium, Burma, Denmark, Italy, Kenya, Kosovo, Kyrgyzstan, Lebanon, Liberia, Mali, Mauritania, Mauritius, Niger, North Macedonia, Oman, Pakistan, Saudi Arabia, Slovakia, Turkey, Uganda, Uzbekistan, Venezuela | Algeria, Chad, Ireland, Luxembourg, Netherlands, Norway, Serbia, South Korea, Sweden, United Arab Emirates |
| D | 3 | Costa Rica, Indonesia, Malaysia, Slovenia | Albania, Argentina, Belarus, Bolivia, Cabo Verde, Canada, Colombia, Comoros, Congo (Kinshasa), Dominican Republic, El Salvador, Eswatini, Germany, Ghana, Guyana, Haiti, Kazakhstan, Lesotho, Libya, Malawi, Mexico, Nigeria, Panama, Russia, Rwanda, Sierra Leone, South Africa, Sudan, Suriname, Switzerland, Thailand, Tunisia, Zambia, Zimbabwe | Andorra, Honduras, Israel, Japan, Monaco, Portugal, Spain, United Kingdom, USA, Vietnam |
|  | 5 | Angola, Eritrea, Latvia, Tajikistan | Liechtenstein, Syria | Burkina Faso |
| C | 4 | Brunei, Central African Republic, China, Equatorial Guinea, Holy See, Tanzania, Yemen | Australia, Djibouti, New Zealand, Singapore, Taiwan |  |

## Discussion

In this paper, we proposed a simple method to analyze the infection curve of COVID-19 and showed that the infection curve can be classified into nine states on the basis of the behavior in the recent past.

In a previous paper [2], it is argued that the apparent steady states of the infection status could be classified into five types: Type I is a steadily increasing infection curve without any wavy behavior, which corresponds to I-1 _1_ or S-2_1_ in the present study, and Type II is a similar state with some number of waves, which corresponds to S-2 _*n*_ (*n* ≥ 2). Type III is a self-organized wavy infection curve which consists of some number of sequence of states I-1, S-2, D-3, S-4, I-5 and I-1. Type IV and V correspond to S-4 and C-4, respectively, in the present classification. The present classification scheme can be used to classify the infection status and the classification provides the basis for further study on the disparity of the outbreak in different countries.

The evolution of infection status shown in Figs. 3 and 4 implies that the key strategy for controlling COVID-19 is to keep *λ* negative and to bring the infection status from D-3 to C-4. In order to realize the strategy, the ingredients in the infection rate *λ* must be specified, which depend on the model employed. Here we exploit the SIQR model [4, 5] which seems to be appropriate to COVID-19 [6]. The infection rate is written as *λ = βS/N* − *q* − *γ*, where the term related to infection process *βS/N* is given by the transmission coefficient *β* which depends on the characteristics of the virus (which might be mutated) and measures reducing the social contacts, and the fraction of susceptible individuals *S* in the population *N* depends on the degree of vaccination on the assumption that the vaccine prevents the transmission of the virus. The quarantine rate *q* ≡ Δ *Q (t)/I(t)* is determined by the quarantine measure, where self-isolation must be included and the regulation is strictly obeyed. The recovery rate *γ* represents the per capita rate at which asymptomatic patients become non-infectious, which cannot be controlled since they cannot be treated. Therefore, policies to make *λ* negative must be either reducing *βS/N* or increasing *q*. In particular, in the step from D-3 to C-4, a measure increasing *q* by quarantining pre-symptomatic and asymptomatic patients effectively is most important and, to make the quarantine measure work, supports for self-isolated people at home are critical as we can see in many countries which succeeded in controlling COVID-19. It has been proved that the quarantine measure is more effective than the lockdown measure in controlling COVID-19 [7].

Vaccination has been started in UK in December, 2020 and is expanding in the whole world, where Israel, UAE, USA and some other countries have managed to speed up the vaccination for COVID-19. However, the infection status analyses as of February 17, 2021 do not show clear difference since the infection is declining in many countries. The effect of the vaccination will be seen as it suppresses the next wave.

## Supporting information

Supplemental data for the main paper

## Data Availability

N.A.

## Author contributions

T. O. conceived and developed the formulation for analysis of the infection status and R. S. conducted analysis of the infection status of 190 countries.

## Additional information

We declare that this research has been conducted independently from any other work and that there are no competing interests.

