## Supplemental data for the main paper for "Classification of the infection status of COVID-19 in 190 countries"

### Supplementary Materials for
### Classification of the infection status of COVID-19 in 190 countries

Reiji Suda[1] and Takashi Odagaki[1,2,*]

[1]Research Institute for Science Education, Inc., Kitaku, Kyoto 603-8346, Japan
[2]Kyushu University, Nishiku, Fukuoka 819-0395, Japan

March 2, 2021

**Abstract**

We present (1) the infection statuses of 24 selected countries from January 23, 2020 to February 17, 2021, (2) classification of the infection statuses of 186 countries as of November 19, 2020 and (3) the infection statuses of 64 selected countries from January 23, 2020 to November 19, 2020.

#### Details of fitting procedure

We first identified maxima, minima and inflection points of the infection curve or the time dependence of the daily confirmed new cases of a given country by careful observation. Using the data of these breaking points, we determined trial piecewise fitting functions from Eqs. (3) $\sim$ (5) and Table 1 of the main paper. Comparing the trial fitting functions with the real data, we adjusted the data for the breaking points until the fitting becomes satisfactory. Since we are interested in qualitative characteristics of the outbreak, we did not use any optimization procedures for the fitting. We plotted the infection rate $\lambda(t)$ as a function of the daily confirmed new cases $\Delta Q(t)$ expressed by the fitting function to get the infection status plot.

#### Infection statuses of 24 countries as of February 17, 2021

We have analyzed the infection curves and the infection status plots of 190 countries from January 23, 2020 to February 17, 2021 in detail. The classification of the infection statuses of these countries is summarized in Table 2 of the main paper, and the infection curves and the infection status plots for 10 typical countries are shown in Fig. 2 of the main paper. In this section, we show results for other 24 selected countries as of February 17, 2021.

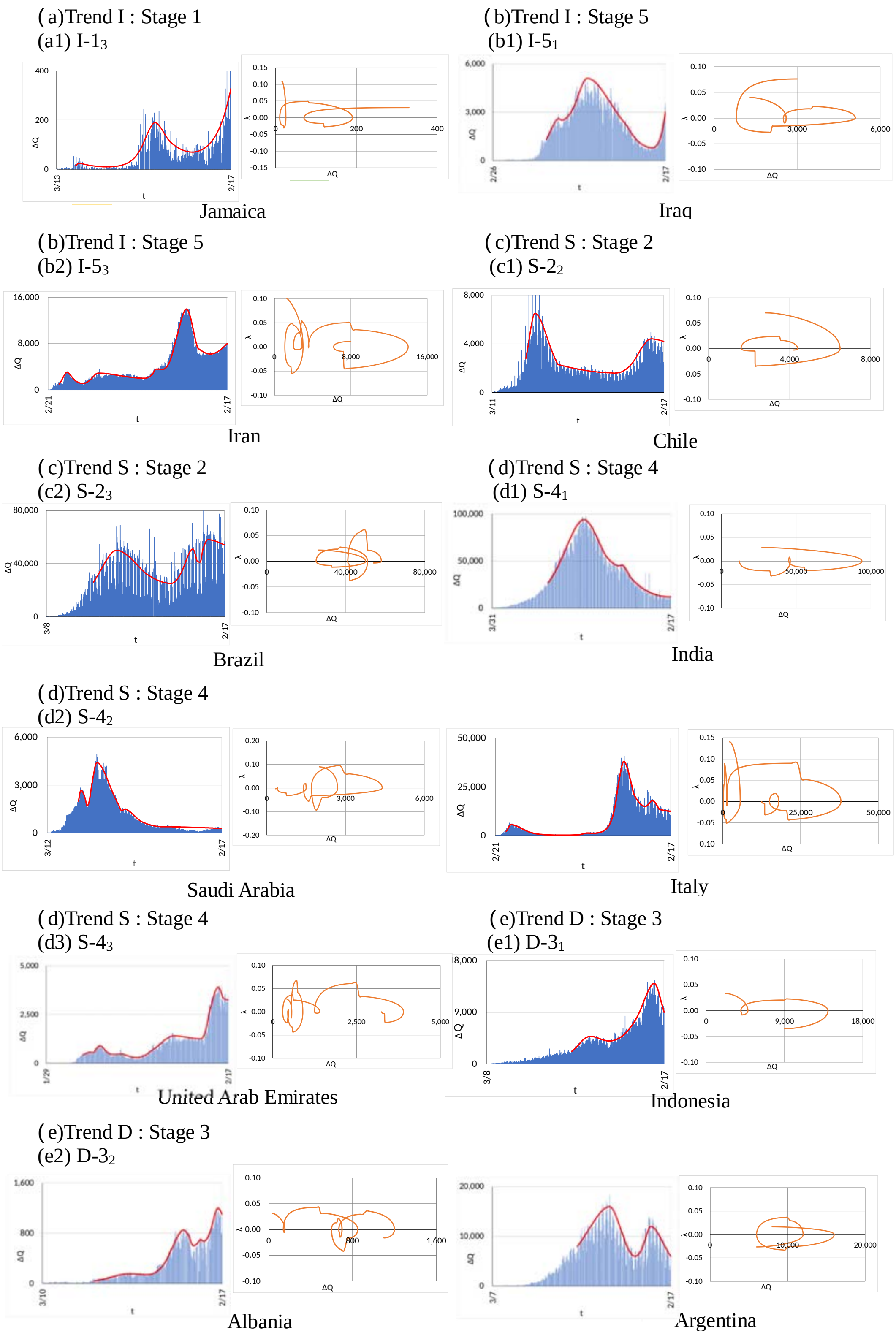


Figure 1: The infections curve and the infection status plot up to February 17, 2021 for countries in Trends I, S and D. In each country, the panel on the left hand side is the infections curve and fitting, and the panel on the right hand side is the infection status plot. (a) shows a country in Trend I and stage 1, (b) shows countries in Trend I and stage 5, (c) shows countries in Trend S and stage 2, (d) shows countries in Trend S and stage 4, and (e) shows countries in Trend D and stage 3. Data from Johns Hopkins University [1].

( e)Trend D : Stage 3
(e2) D-$3_2$

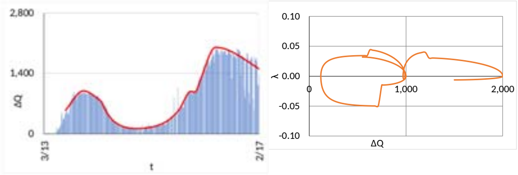
Belarus

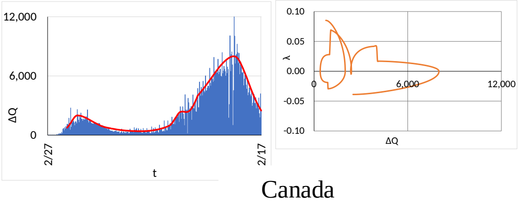
Canada

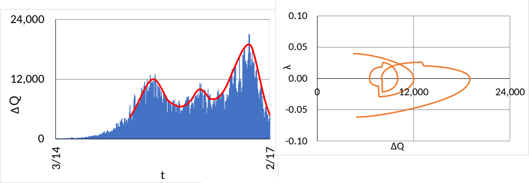
Colombia

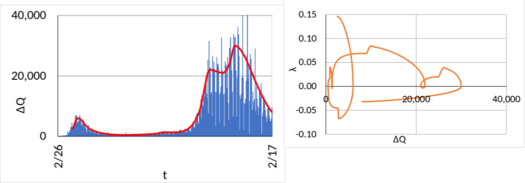
Germany

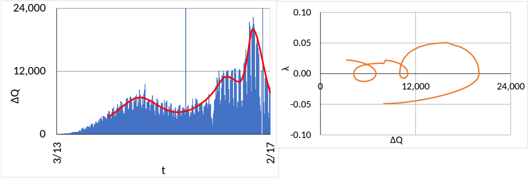
Mexico

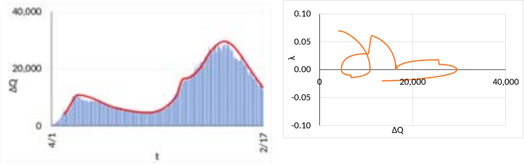
Russia

(e2) D-$3_2$

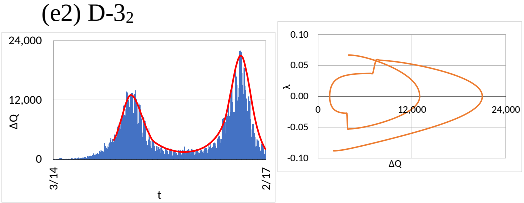
South Africa

(e3) D-$3_3$

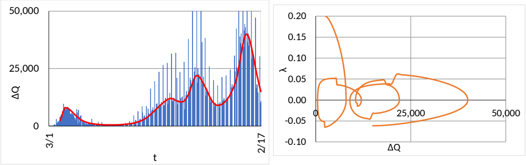
Spain

(e3) D-$3_3$

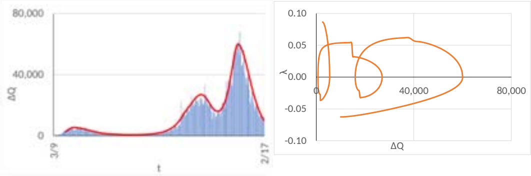
United Kingdom

( f)Trend D : Stage 5
(f1) D-$5_1$

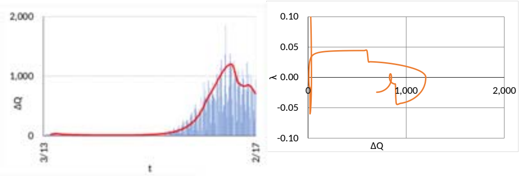
Latvia

( g)Trend C : Stage 4
(g1) C-$4_2$

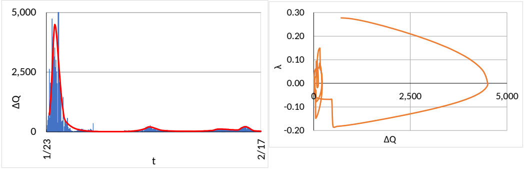
China

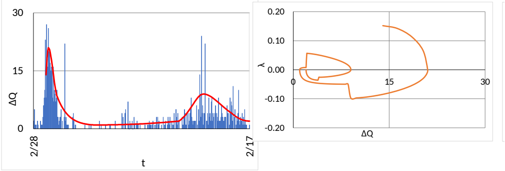
Taiwan

Figure 1: (Continued) The infections curve and the infection status plot up to February 17, 2021 for countries in Trends D and C. In each country, the panel on the left hand side is the infections curve and fitting, and the panel on the right hand side is the infection status plot. (e) shows countries in Trend D and stage 3. (f) shows a country in Trend D and stage 5, and (g) shows countries in Trend C, Data from Johns Hopkins University [1].

### Classification of the infection status of 186 countries as of November 19, 2020

The infection status of each countries changes drastically in time. Table 1 shows the classification of the infection status of the COVID-19 outbreaks in 186 countries as of November 19, 2020, which can be compared with Table 2 of the main paper. We noticed significant changes of the infection status in three months.

Table 1: Classification of the infection status of COVID-19 in 186 countries as of November 19, 2020. The first and second columns are Trend and Stage, respectively. The infection status is identified by Trend, Stage and Wave and the status is labeled as T-$k_n$, where T denotes a trend (increasing (I), stationary (S), decreasing (D) and converged(C)), $k$ and $n$ represent a stage and the current wave. Countries are listed in alphabetical order in each state.

| T | S | Countries | | |
|---|---|---|---|---|
| | | Wave 1 | Wave 2 | Wave 3 $\sim$ |
| I | 1 | Botswana, Georgia, Hungary, Jordan, Ukraine | Albania, Azerbaijan, Indonesia, Kosovo, Latvia, Pakistan, Portugal, San Marino, Somalia, Sudan, Syria, Uganda, West Bank and Gaza | Algeria, Iran, Japan, Panama, Serbia, South Korea, Turkey, USA |
| | 3 | Belize, Greece, Togo, Uruguay | Belarus, Canada, Estonia, Russia | Moldova |
| | 5 | Afghanistan, Bangladesh, Benin, Brazil, Burma, Burundi, Comoros, Congo (Brazzaville), Congo (Kinshasa), Costa Rica, Egypt, El Salvador, Ghana, Jamaica, Kazakhstan, Liberia, Mali, Paraguay, Rwanda, South Africa, Trinidad and Tobago, Zimbabwe | | |
| S | 0 | Antigua and Barbuda, Barbados, Bhutan, Cambodia, Dominica, Eritrea, Fiji, Grenada, Holy See, Laos, Liechtenstein, Mongolia, Saint Kitts and Nevis, Saint Lucia, Saint Vincent and the Grenadines, Seychelles, Western Sahara | | |
| | 2 | Ecuador, Guinea, Morocco, Poland, Sri Lanka, Tunisia | Austria, Bosnia and Herzegovina, Bulgaria, Croatia, Cuba, Cyprus, Finland, Germany, Italy, Kenya, Kyrgyzstan, Libya, Lithuania, Malaysia, Malta, North Macedonia, Norway, United Kingdom | Denmark, Lebanon, Luxembourg, Montenegro, Romania, Sweden |
| | 4 | Bahamas, Cameroon, Chile, Dominican Republic, Eswatini, Guatemala, Guyana, India, Iraq, Namibia, Nigeria, Qatar, Slovenia, South Sudan, Tajikistan, Venezuela | Maldives, Peru, Saudi Arabia, Senegal, Uzbekistan | Ethiopia, Israel |
| D | 3 | Angola, Argentina, Cabo Verde, Cote d'Ivoire, Czech, Honduras, Nicaragua, Slovakia, Zambia | Andorra, Armenia, Bahrain, Belgium, Burkina Faso, Chad, Colombia, France, Iceland, Ireland, Lesotho, Mexico, Monaco, Mozambique, Nepal, Netherlands, Oman, Spain, Switzerland, United Arab Emirates | Kuwait, Philippines |
| | 5 | Sierra Leone | | |
| C | 4 | Bolivia, Brunei, Central African Republic, China, Equatorial Guinea, Gabon, Gambia, Guinea-Bissau, Haiti, Madagascar, Malawi, Mauritania, Mauritius, Niger, Papua New Guinea, Sao Tome and Principe, Suriname, Taiwan, Tanzania, Thailand, Timor-Leste, Yemen | Australia, Djibouti, New Zealand, Singapore, Vietnam | |

### Infection curves and infection status plots of 64 countries from 23 January, 2020 to November 19, 2020

We analyzed the daily confirmed new cases $\Delta Q(t)$ in 186 countries from January 23 to November 19, 2020 provided by Johns Hopkins Coronavirus Resource Center [1]. We show in Figs. 2 $\sim$ 5 the infection curves and the infection status plots for 64 selected countries. Note that the scale of axes are different depending on the country.

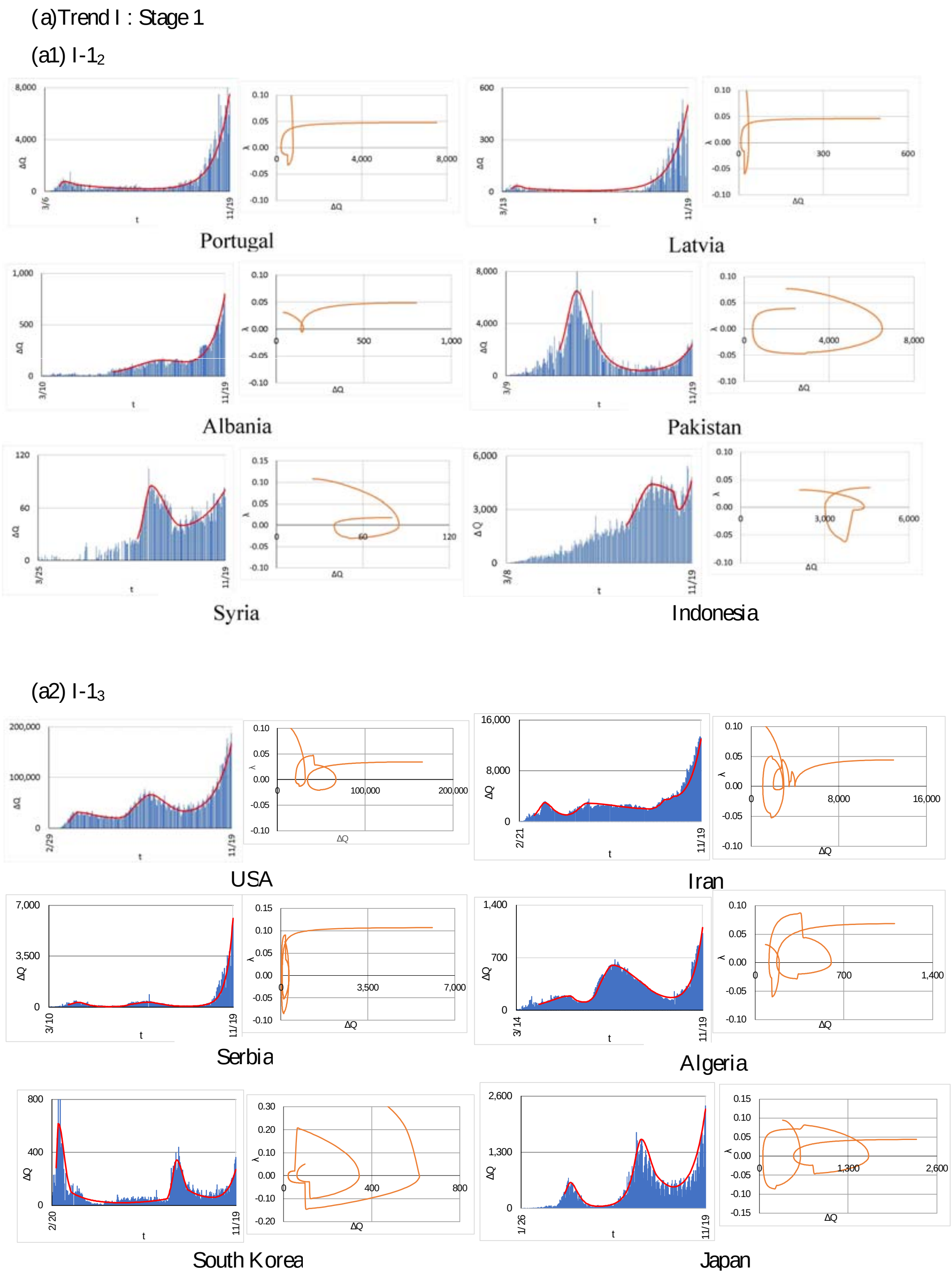


Figure 2: The infection curve and the infection status plot up to November 19, 2020 for countries in Trend I. In each country, the panel on the left hand side is the infection curve and fitting, and the panel on the right hand side is the infection status plot. (a) shows the countries in Trend I and stage 1, where (a1) and (a2) are the countries in the second and the third waves, respectively. Data from Johns Hopkins University [1].

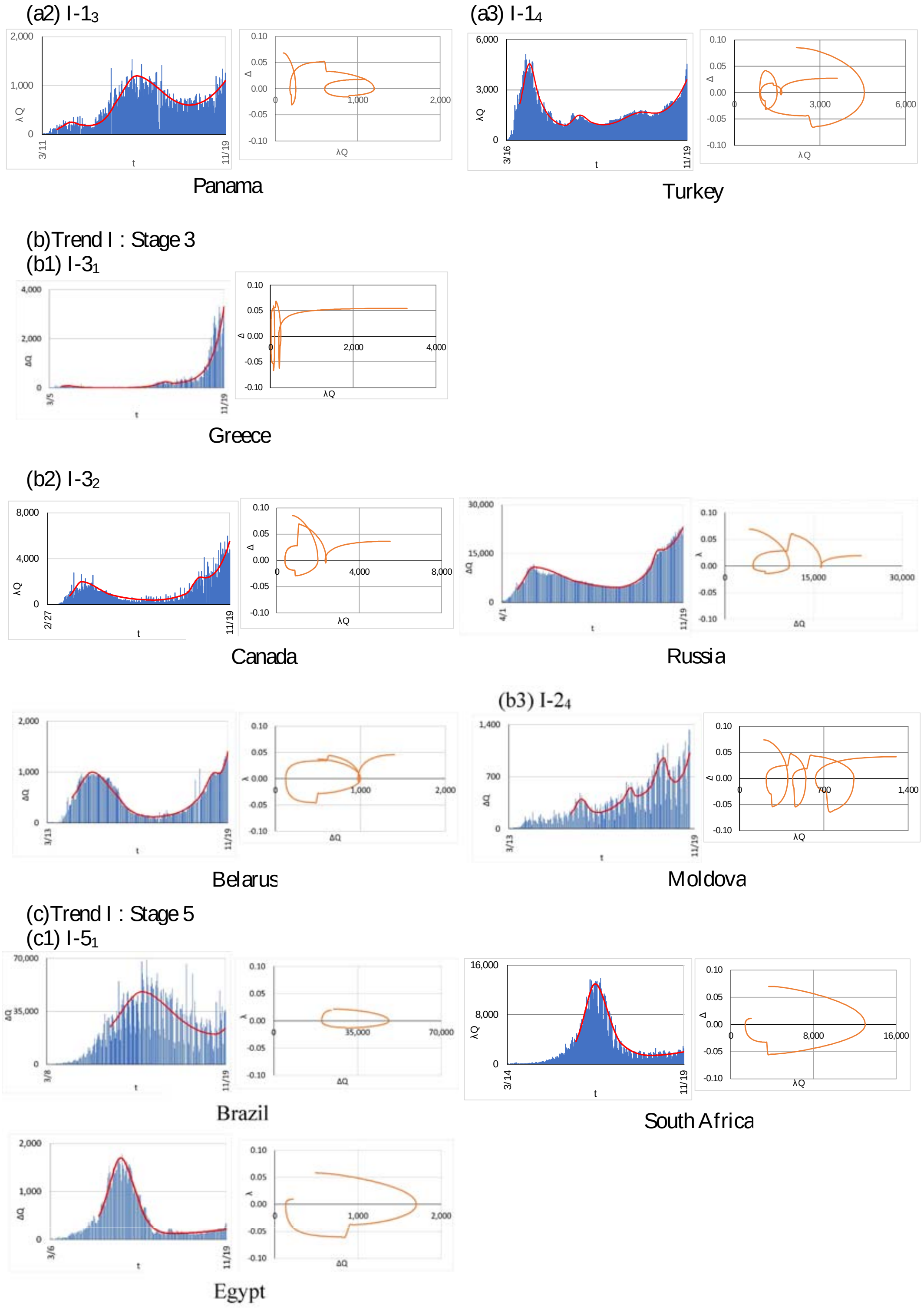


Figure 2: (Continued) The infection curve and the infection status plot up to November 19, 2020 for countries in Trend I. In each country, the panel on the left hand side is the infection curve and fitting, and the panel on the right hand side is the infection status plot. (a2) and (a3) show the countries at stage 1 in the third and the fourth waves. (b) shows the countries in Trend I and stage 3, where (b1), (b2) and (b3) are the countries in the first, the second and the fourth waves, respectively. (c) shows countries in Trend I and stage 5. Data from Johns Hopkins University [1].

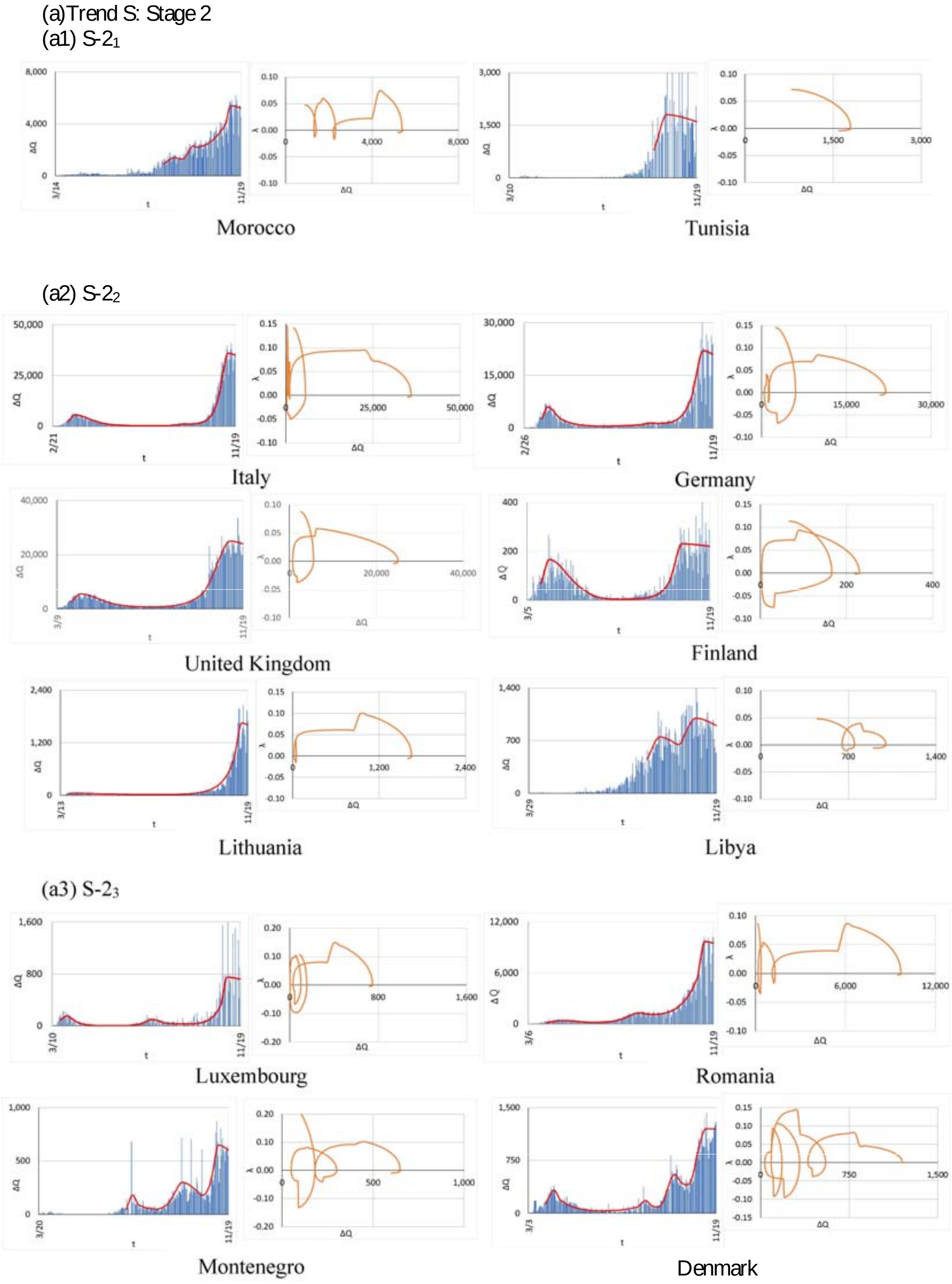


Figure 3: The infection curve and the infection status plot up to November 19, 2020 for countries in Trend S. In each country, the panel on the left hand side is the infection curve and fitting, and the panel on the right hand side is the infection status plot. (a) shows the countries in Trend S and stage 2, where (a1), (a2) and (a3) are the countries in the first, the second and the third waves, respectively. Data from Johns Hopkins University [1].

(a)Trend S: Stage 2

(a3) S-$2_3$

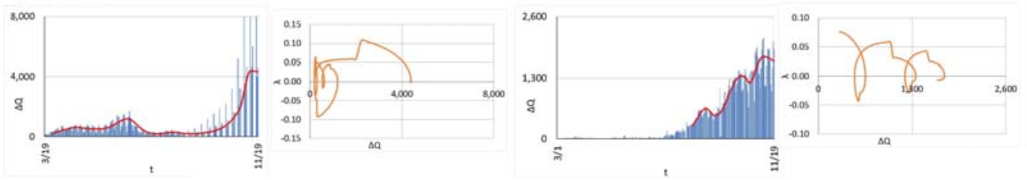

Sweden Lebanon

(b)Trend S: Stage 4

(b1) S-$4_1$

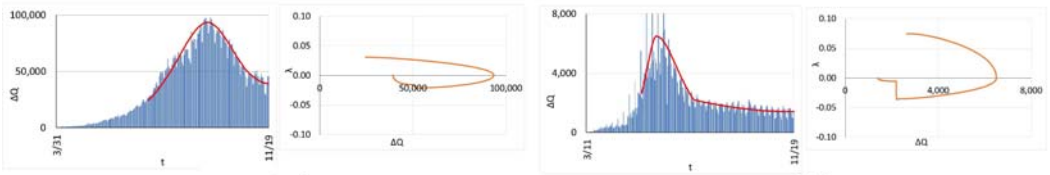

India Chile

(b2) S-$4_2$

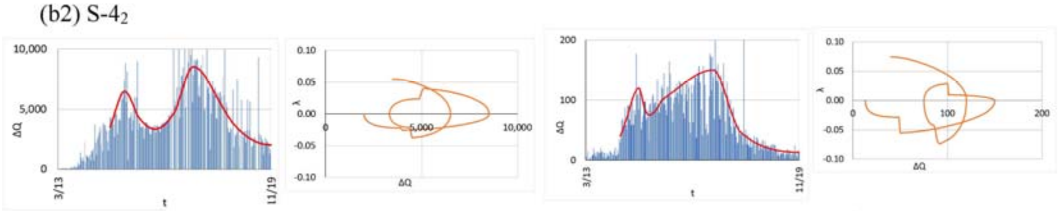

Peru Senegal

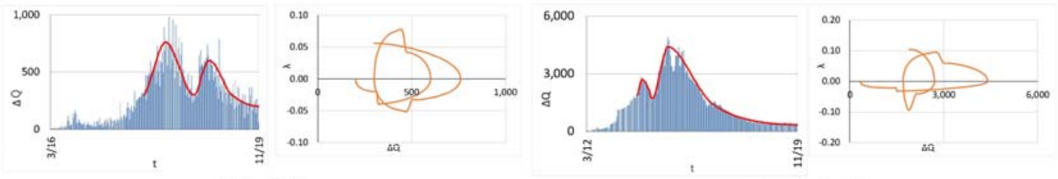

Uzbekistan Saudi Arabia

(b3) S-$4_3$

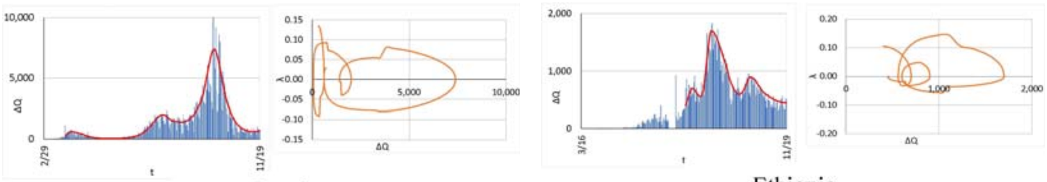

Israel Ethiopia

Figure 3: (Continued) The infection curve and the infection status plot up to November 19, 2020 for countries in Trend S. In each country, the panel on the left hand side is the infection curve and fitting, and the panel on the right hand side is the infection status plot. (a3) shows the countries in Trend S and stage 2 in the third wave. (b) shows the countries in Trend S and stage 4, where (b1), (b2) and (b3) are the countries in the first, the second and the third waves, respectively. Data from Johns Hopkins University [1].

(a)Trend D: Stage 3

(a1) D-$3_1$

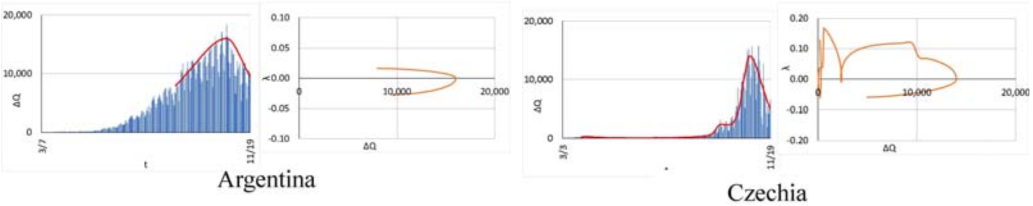


(a2) D-$3_2$

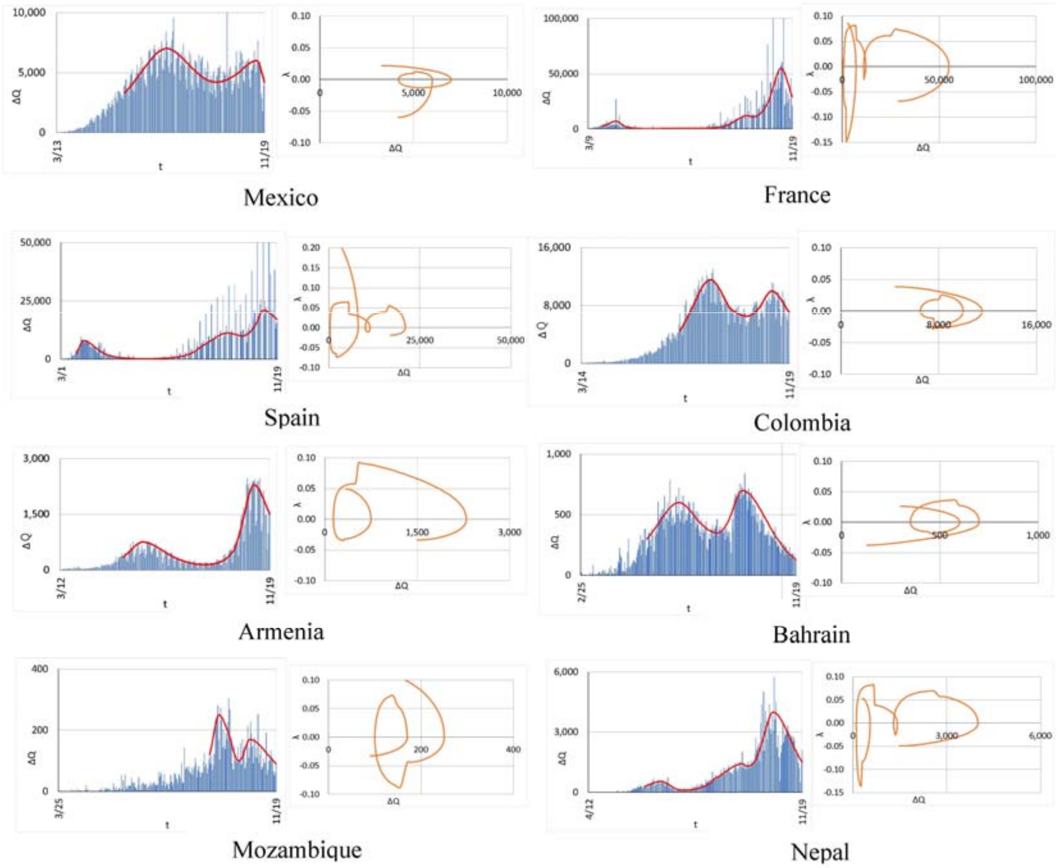


(a3) D-$3_4$

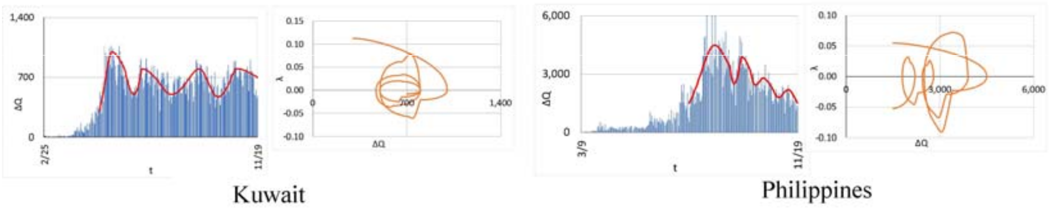


Figure 4: The infection curve and the infection status plot up to November 19, 2020 for countries in Trend D. In each country, the panel on the left hand side is the infection curve and fitting, and the panel on the right hand side is the infection status plot. (a) shows the countries in Trend D and stage 3, where (a1), (a2) and (a3) are the countries in the first, the second and the fourth waves, respectively. Data from Johns Hopkins University [1].

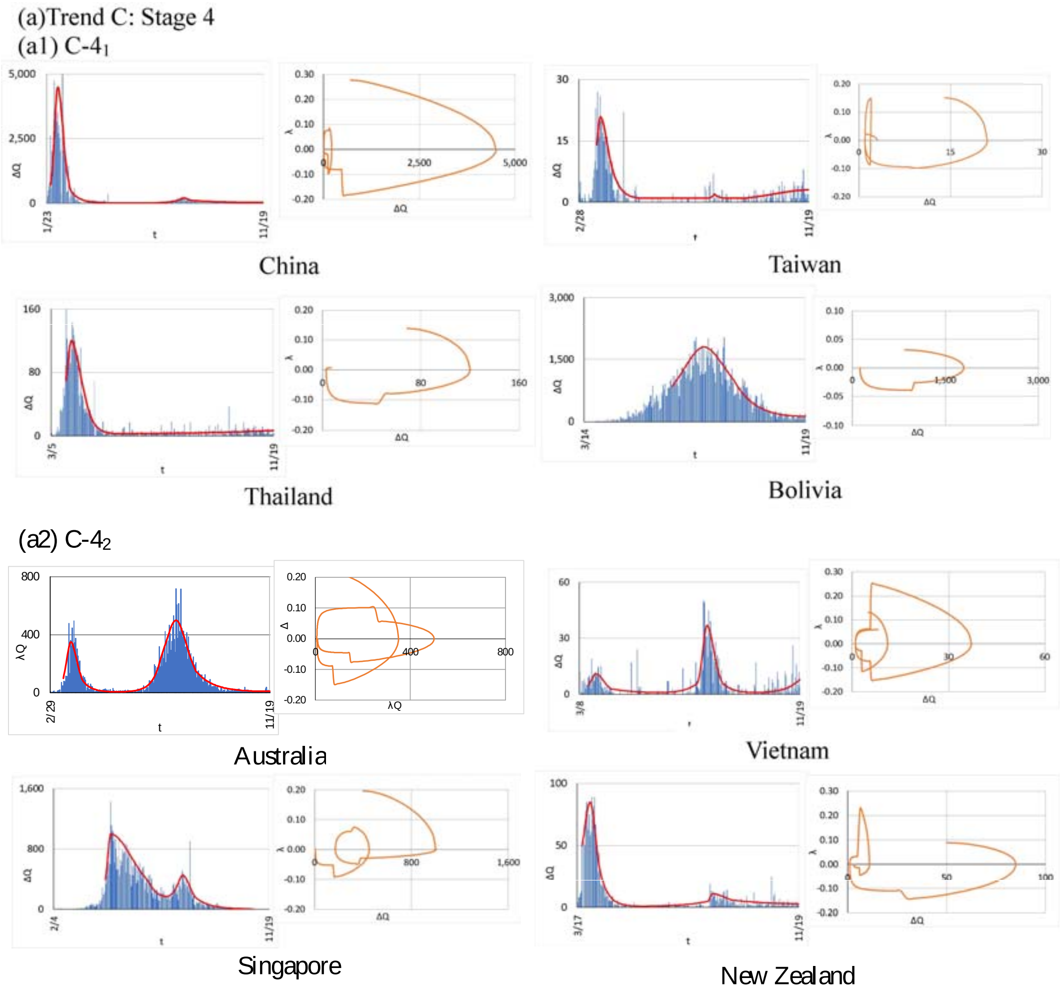


Figure 5: The infection curve and the infection status plot up to November 19, 2020 for the countries in Trend C and stage 4. In each country, the panel on the left hand side is the infection curve and fitting, and the panel on the right hand side is the infection status plot. (a1) and (a2) are the countries in the first and the second waves, respectively. Data from Johns Hopkins University [1].

### References

[1] Coronavirus Resource Center, Johns Hopkins University; https://coronavirus.jhu.edu/

### Author contributions

T. O. conceived and developed the formulation for analysis of the infection status and R. S. conducted analysis of the infection status of 190 countries.

### Additional information

We declare that this research has been conducted independently from any other work and that there are no competing interests.
